# A mathematical modelling framework for the “stop when you feel better” approach to antibiotic prescribing

**DOI:** 10.64898/2026.08.03.26359575

**Authors:** Alice Ledda, Stephanie Evans, Luca Ferretti, Laurence Roope, Diane Pople, Jack Pollard, Aleksandra J. Borek, Koen B. Pouwels, Sarah Tonkin-Crine, A. Sarah Walker, Julie V. Robotham

## Abstract

To prevent antimicrobial resistance (AMR), antibiotic courses are traditionally prescribed to completion, overlooking their collateral impact on the commensal microbiota. We developed a deterministic within-host model linking pathogen growth, immune responses, antibiotic action, and commensal dynamics to explore how treatment duration influences resistance development. Simulations across community-acquired infection parameters showed that 71% of cases successfully treated with a standard 7-day regimen could also be cured with shorter “Stop When Better” (SWB) courses, where treatment cessation was triggered by a predefined threshold in pathogen abundance, and 97% by SWB plus 1 day. Compared with standard treatment, SWB+1 approaches reduced overall antibiotic exposure and resistance emergence in commensal niches while preserving efficacy. While this threshold-based definition of “feeling better” offers a clear theoretical framework, it would require translation into clinical cues. Therefore, although these findings suggest that response-based antibiotic stopping rules could inform future adaptive prescribing strategies, further empirical validation is essential.

## Introduction

Taking the entire course of antibiotic as prescribed and not stopping any earlier to avoid unduly causing resistance in the pathogen is a dogma dating back to Sir Alexander Fleming ***Fleming (1947)***. He noted that if the pathogen was not completely wiped out during treatment it could develop ways of avoiding being killed by it, something that we now term resistance ***Murray et al. (2022)***. For years patients were educated to continue to take the antibiotic for the full prescribed duration, even if they feel better prior to this, with the idea that this will minimise resistance development and ensure clearance of infection ***CDC (2025)***; ***PHE (2025)***.

This dogma, and standard practice, focuses on the pathogen causing the infection, aiming at avoiding incomplete pathogen clearance which could lead to relapse of symptoms; however, as a bacterial host, the human body is a complex environment with extensive cross-interactions across many niches, each with its own collection of commensal organisms. If a patient is prescribed a course of antibiotics, the drug will affect not only the pathogen but the entire microbiome Rel- man and Lipsitch (2018); ***Peto et al. (2025)***. During an antibiotic course, multiple bacterial populations will be exposed, with the potential for resistance emergence and expansion, referred to as “bystander resistance” ***Tedijanto et al. (2018)***; ***Morley et al. (2019)***. Impacts like niche depletion and resistance emergence in commensal bacteria could have more serious consequences for the patient over a medium/long timescale (months or years following antibiotic treatment) than any effects on the pathogen ***Llewelyn et al. (2017)***.

Mathematical modeling of antimicrobial resistance (AMR) has traditionally focused on the target pathogen population, considering how treatment duration, dosing, and combination therapy influence resistance evolution within hosts to minimize resistance across populations ***Bonhoeffer et al. (1997)***; ***Levin et al. (1997)***; ***Austin et al. (1999)***; ***Lehtinen et al. (2017)***. More recently, modeling efforts have focused on bystander selection, explicitly taking into account that antibiotics exert selective pressure on non-target commensal or colonizing bacteria that are merely exposed during therapy ***Tedijanto et al. (2018)***; ***Morley et al. (2019)***. In parallel, observational studies have been studying the effect of different antibiotics on the patient’s microbiota, using heterogeneous approaches, which complicates interpretation of the findings ***Malhotra-Kumar et al. (2007)***; Deth-lefsen et al. (2008); ***D’humières et al. (2024)***; ***Chu et al. (2025)***.

Rethinking the “take the whole antibiotic course as prescribed” dogma ***Llewelyn et al. (2017)*** and letting patients stop antibiotics when they “feel better” might be an approach to achieve shorter yet still effective antibiotic courses for relatively mild-modest infections treated in the community e.g. respiratory tract infections, which are often self-limiting or caused by viruses. Shorter antibiotic courses could successfully clear the causative pathogen while minimising the commensals’ antibiotic exposure, lowering the chance of developing resistance without significantly impacting the risk of relapse or severe infection-related outcomes. Whilst not prescribing at all may be ideal in these scenarios, this can be challenging given diagnostic uncertainty and patient pressure.

While “stopping when better” could in theory reduce antibiotic exposure and associated side-effects, the intrinsic difficulty is defining what ‘feeling better’ means, as it is key for the success of this approach. If the pathogen is not cleared by a shorter course of antibiotics, the infection might relapse, needing a further antibiotic course, leading to a longer total duration of antibiotics, effectively increasing the overall chances for commensals to develop resistance.

To explore the consequences of antibiotic treatment not only on the pathogen but also on the commensal microbiome, we built a mathematical model of the system. We use this model to demonstrate the possibility of “stopping when better” (SWB) being more effective for certain pathogen and antibiotic characteristics.

## Methods and Results

### A model for the within-patient effect of antibiotic treatment

We developed a mathematical model to describe the processes occurring at a patient-level during a mild-moderate bacterial infection and its subsequent antibiotic treatment in the community. This framework allows extrapolation to viral infections, for which antibiotics are not needed and where clinical recovery would be expected to be independent of treatment duration. The model is described by a set of deterministic ordinary differential equations and is structured into nested blocks that can link together to expand the scope of the within-patient interaction. The basic and most restricted model block focuses on the interaction between the pathogen, the patient’s immune system and the antibiotic, the larger model block has the capability to include the interaction with commensals, divided into separate niches. The final model block expands to the interaction with the environment. A sketch of the model is given in figure 1. The wide parameter space we explore (see below) covers bacteria with different growth rates and dynamics.

**Figure 1.**
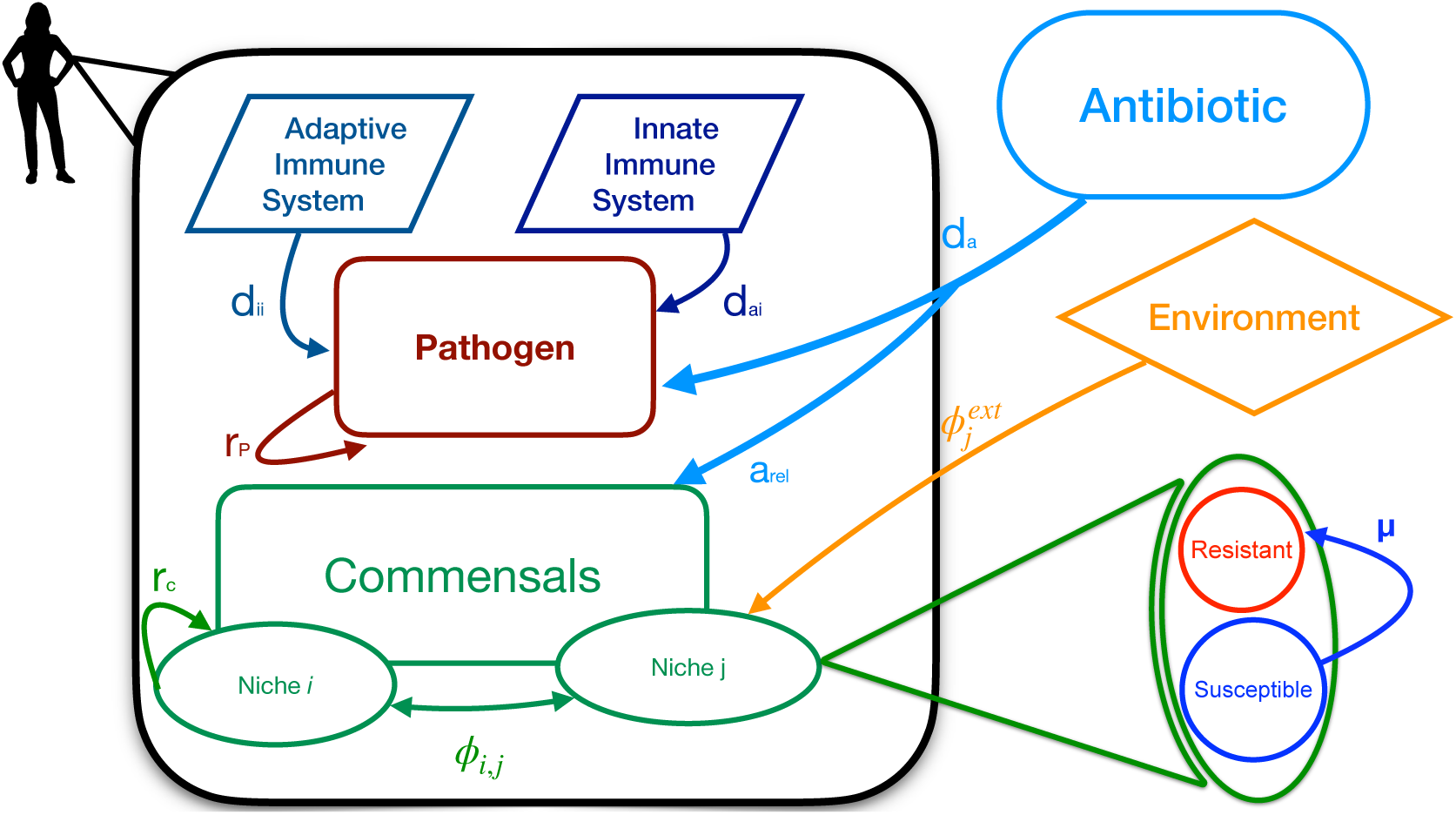
Sketch of the model. The model focuses on the within-patient dynamic of antibiotic treatment (black box). Within the patient there is a pathogen population (whose replication rate is *r_p_*) and a commensal population divided into several niches (each having a niche specific replication rate *r_c_*). The adaptive and innate immune system attack the pathogen with a killing rate *d_ii_* and *d_ai_* respectively. Each commensal niche can have both susceptible and resistant bacteria and susceptible bacteria develop resistance (during antibiotic exposure) at a rate *μ*. Flux of bacteria between different niches happens at a rate *φ_i_*_,*j*_. The pathogen and the commensals niches do not interact except for the effect of the antibiotic, which acts on the pathogen with a killing rate *d_a_* and on the commensals with a multiplicative killing rate *a_rel_* (niche specific). Commensals niches can import bacterial from the environment at a rate *φ^ext^_i_*. For more details on the dynamics see the section “Description of the model” and supplementary.

In our model, when a patient is infected by an external bacterial pathogen, the pathogen establishes a new niche, with no resource restriction, where it grows exponentially (figure 2). During this time the patient’s innate immune system is working to protect the patient, de facto providing a reduction on the pathogen’s growth rate. When the pathogen abundance reaches a specific threshold (called *“noticeable threshold”*) the patient starts experiencing symptoms, triggering both the antibiotic treatment and the adaptive immune system. Both result in a further reduction of the growth rate of the pathogen. The model can be evaluated in different prescription scenarios, namely where the antibiotic is prescribed for a fixed amount of days or where the antibiotic stopping is triggered by the pathogen reaching a pre-determined threshold, called *“recovery threshold”* (see figure 2A). We used this threshold as a proxy for main symptoms (i.e. fever) being resolved.

**Figure 2.**
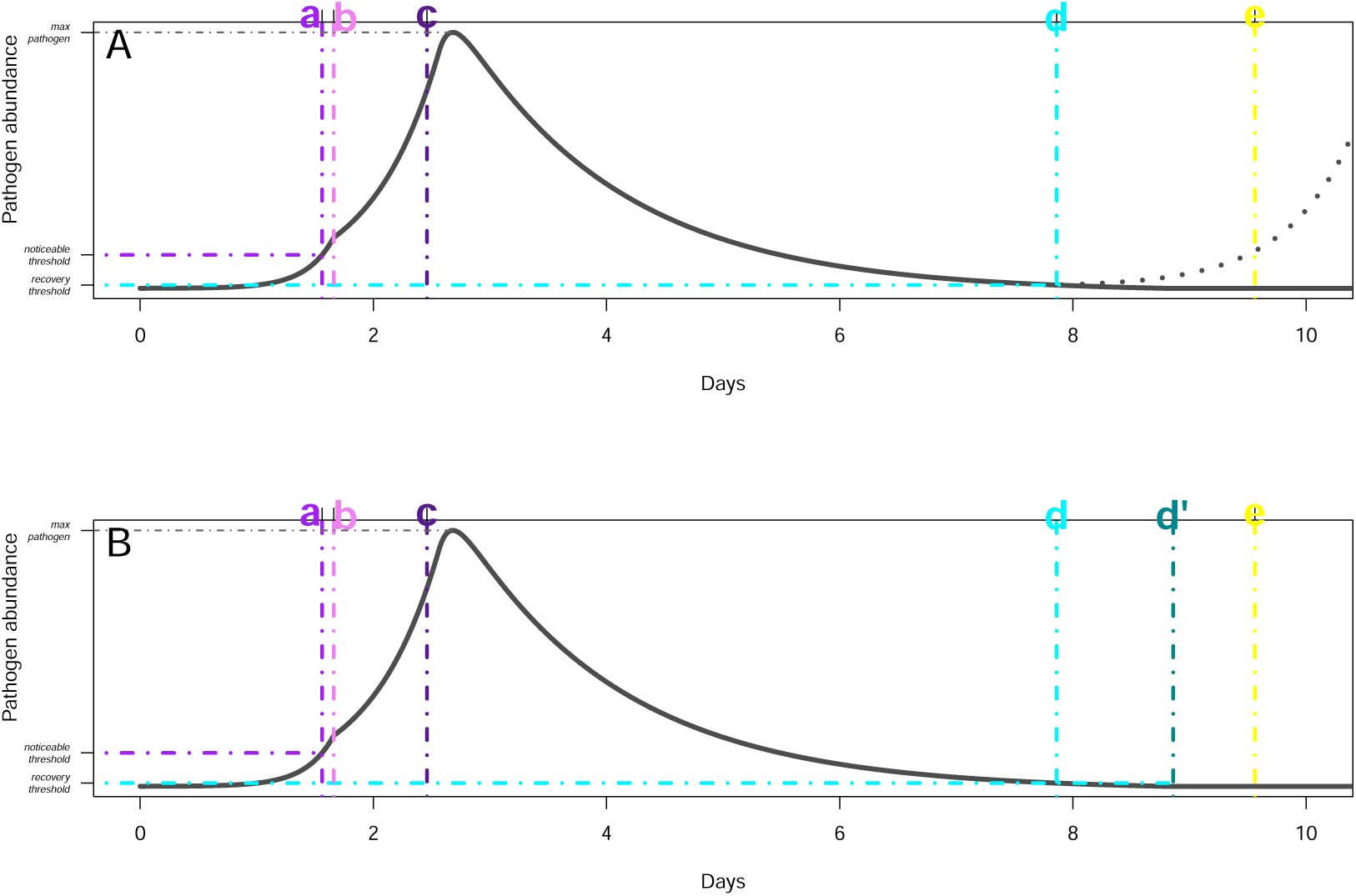
General pathogen dynamics. **A-SWB:** The pathogen enters the patient at time 0 and starts to grow (black line depicts pathogen abundance) until it reaches the noticeable threshold, indicated with the purple horizontal line. The timing at which it reaches the noticeable threshold is time “a”, indicated by the purple vertical line. This triggers both the adaptive immune system which starts at time “b” indicated by the pink vertical line and the antibiotic treatment, which starts one day after the threshold is reached, at time “c”, shown by the dark purple line. The antibiotic does not reach its maximum killing rate immediately, but its killing rate grows linearly until saturation. The pathogen abundance decreases until it reaches the “recovery threshold” (horizontal light blue line) where the treatment is stopped in the SWB regime (the timing is shown by the vertical light blue line “d”). In the standard treatment scenario the treatment goes on for a full 7 days after the beginning of the treatment, displayed by the yellow vertical line “e”, which is 7 days (the length of the standard treatment course) from the dark purple line “c”. **B-SWB+1:** the same simulation in the SWB+1 scenario. In this case the antibiotic treatment is stopped one day after (dark blue line “d’”) the pathogen concentration reaches the “recovery threshold” (light blue line “d”). Parameters for this image are *r_p_* = 6.63, *d_a_* = 2.45, *d_ai_* = 2.45, *d_ii_* = 2.45, noticeable threshold= 678, recovery threshold= 0.01 (times the noticeable threshold). In this case SWB is not sufficient to ensure that the infection is wiped out, hence, once the antibiotic is stopped, the infection rebunds (dotted line on the left). SWB+1 leads to a complete recovery.

In the default scenario the antibiotic is stopped after a *“standard duration”* (7 days), while the adaptive immune system is never switched off (figure 2A) within the simulation period.

The antibiotic (both in terms of treatment duration and antibiotic killing rate) also affects the niches of commensals. Each niche is characterised by its size (carrying capacity) and growth rate and a multiplicative parameter that enhances or dampens the antibiotic killing rate (figure 1).

Different niches can exchange bacteria at different rates and each niche can have exchanges with the environment.

The model is implemented in a set of ordinary differential equations with no stochasticity, except in the interaction with the environment. Therefore, once the parameters are set, the dynamic of the pathogen infection is fully determined (as in the examples shown in figure 2). When investigating the combination od parameters (region of parameter space) that presented an interesting dynamics for this problem, a very strong constraint is given by the kind of infections we aim to study. We focus on non-chronic infections that are attended to in primary care. As such we require that they become acute (pathogen abundance reaches *noticeable threshold*) in a finite and relatively short time (around 2 weeks) from infection. Therefore, for example, only pathogen replication rates that satisfy this constraint are interesting for simulations. Pathogen replication rates that would make the pathogen reach the noticeable threshold in 5 minutes or in two years are not interesting for the purposes of this study, therefore we will not include them in the simulations.

Once the noticeable threshold is fixed, whether the infection becomes acute within the observation time is entirely determined by the values of the pathogen growth rate and innate immune system killing rate (figure 3 for classification and figure 4 for examples of the pathogen dynamics). Simulations where the pathogen abundance never reaches a noticeable threshold are categorised as *“failed infections”* (figure 3). This describes the cases where the immune system neutralizes the pathogen exposure before it can establish infection and cause symptoms (figure 4 top row, A & E).

**Figure 3.**
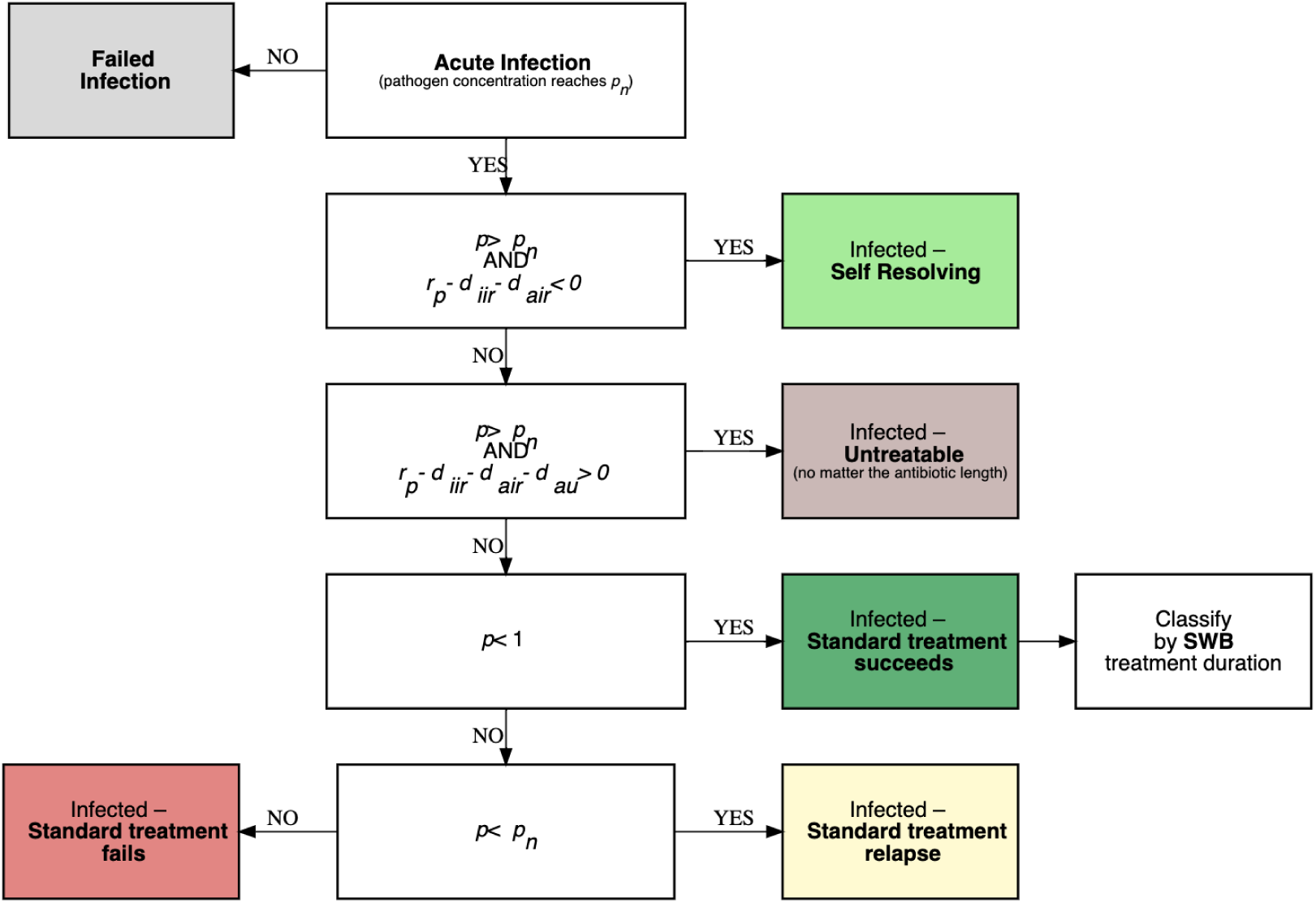
Workflow of the classification of the infection scenarios. If the pathogen abundance does not reach the noticeable threshold *p_n_* during the first 14 days of the simulation we classify the scenario as *“failed infection”* (see figure 4 A,E upper line). If the pathogen abundance reaches *p_n_* during the first 14 days of the simulation, we first classify the infection as *“Infected - Self resolving”* (figure 4 B,F second line) and *“Infected - Untreatable”* (figure 4 C,G third line) depending on the strength of the immune system killing rate compared to the pathogen replication rate. These classifications rely only on the simulation parameters. The infections that are not in either of these previous categories are treatable (figure 4 D,H bottom line) and are further classified depending on the simulation outcomes in the standard treatment regime as “Infected - Treatable - Standard treatment fails”, “Infected - Treatable - Standard treatment relapse” or “Infected - Treatable - standard treatment succeeds”, depending on the pathogen concentration at the end of the simulation, namely if there is any pathogen left, if the successive treatment can send it below p_n_, locking it in an endless cycle of relapses, or if the pathogen keeps growing indefinitely. The scenarios in which standard treatment succeeds are then classified depending on the treatment duration in the SWB (and SWB+1) scenario.

**Figure 4.**
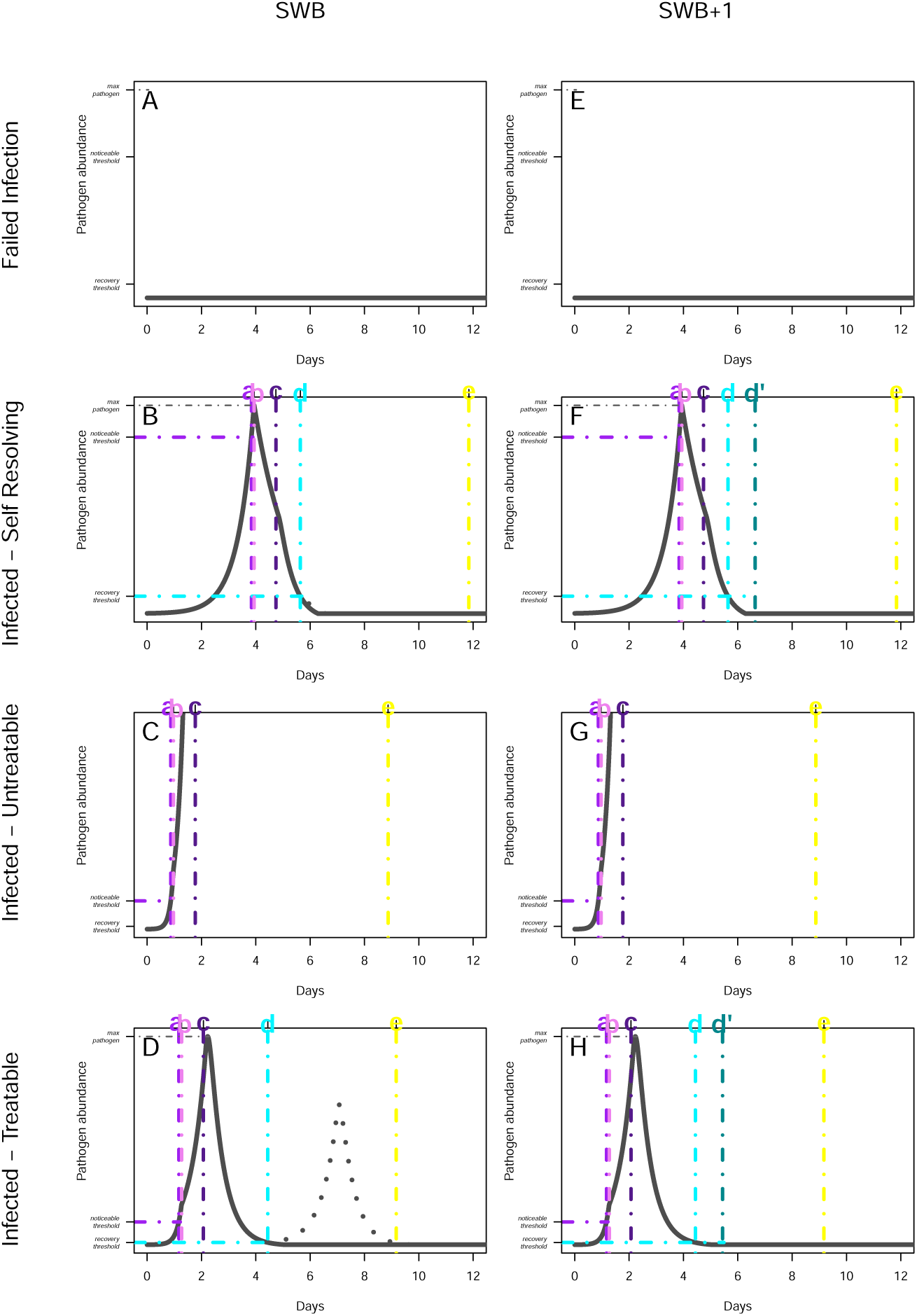
First level classification of infection outcomes. Simulation examples of the first level classification outcomes. The two columns show the simulations made in the **SWB (left)** and **SWB+1 (right)** treatment scenarios. **Failed Infection (A** & **E - top line).** The pathogen replication rate is not strong enough and the pathogen abundance fails to reach the noticeable threshold during the first 14 days of the simulation time. **Infected - Self resolving (B** & **F - second line).** The infection becomes acute but the joint killing power of the innate and adaptive immune system is enough to bring the pathogen abundance down to the recovery threshold without antibiotic treatment. **Infected - Untreatable (C** & **G - third line).** The joining killing rate of the innate and adaptive immune system and the antibiotic killing rate are not enough to curb the pathogen’s growth - no matter the length of the antibiotic treatment. **Infected - Treatable (D** & **H - bottom line).** The combined killing rate of the antibiotic and the innate and adaptive immune system are enough to curb the pathogen spread, but treatment success depends on treatment length. In this specific case SWB treatment (left) is not long enough and is followed by a relapse (dotted line), while in SWB+1 there is no relapse.

If the infection does become acute (and hence shows symptoms), based on the values of all the rates involved in the dynamics, we have 3 possible outcomes: *self-resolving*, *treatable* and *untreatable* (figure 3 and figure 4). For a *self-resolving* infection, no antibiotic treatment is needed as the combined effects of the innate and adaptive immune system will eventually clear the pathogen and therefore symptoms will resolve without antibiotics (figure 4 second row, B & F). In *untreatable* infections, the pathogen will keep growing irrespective of the antibiotic course duration, as the combined action of the antibiotic and the innate and adaptive immune systems is not sufficient to stop the pathogen proliferating (figure 4 third row, C & G). A *treatable* infection is one that, while not self-subsiding, subsides with antibiotic treatment (figure 4 bottom row, D & H). The mathematical definition of this classification is given in supplementary 1 and in the flowchart in figure 3.

Whether a treatable infection fully subsides or relapses depends on the duration of the antibiotic course (figure 3). For a fixed *“standard”* length of antibiotic treatment, the treatable infection, in the short term, can have three possible outcomes: standard treatment *succeeds*, *fails* or (infection) *relapses* (figure 3). Treatment outcomes are determined by final pathogen abundance at simulation endpoint. Successful treatment requires complete pathogen elimination; any detectable pathogen presence is classified as treatment failure. Relapsing infections are identified as those where pathogen levels initially drop below the *“recovery threshold”* but subsequently rebound above the *“noticeable threshold”* during the 28-day observation period, triggering an additional antibiotic course. Distinguishing between *“treatment failure”* and *“relapsing infection”* is crucial because each scenario results in different antibiotic exposure for commensal bacteria. The mathematical details and operative definitions of the outcomes are detailed in supplementary 1.

### Classification of the model commensals related

The model explicitly describes the dynamics within the commensal niches. The parameters that describe the commensals dynamics in our model are the commensal replication rate, the niche size and the niche-relative antibiotic effect. The commensal replication rate takes into account the fact that the microbiome comprises different combinations of bacterial species in each niche and therefore in each niche the replication rate will be different. The niche size is important because the smaller the niche, the easier it is for it to become fully resistant even after a very brief exposure to antibiotics. The realism of this simplified model is enhanced by accounting for the pharmacodynamics of antibiotic treatment and specifically the fact that antibiotics do not act with the same intensity on all the niches within the human body. In our simplified model, the antibiotic effect on commensals is simulated as a multiplicative constant (relative antibiotic effect) to the antibiotic killing rate on the pathogen.

To simplify we used 3 possible classifications for the resistance outcomes of each niche after the antibiotic treatment. Each niche starts with 0.1% of its population being resistant. If the percentage of resistant bacteria in the niche at the end of the simulation does not exceed 1% of its population we classify the niche as having *“baseline resistance”*. If, on the other hand, the resistant population exceeds 99% of the niche population, we classify the niche as *“fully resistant”*. If the resistant fraction of the niche population is anywhere between 1% and 99% we classify the niche as *“partially resistant”*.

### General Model Outcome - pathogen impact

To gain a better understanding of the regions of the parameters space where an infection treatable in primary care arises, we ran a series of simulations on a latin hypercube covering a combination of values of the parameters (see supplementary table 5 for details). Each combination of specific values of the parameters will be referred to as a “simulation scenario” for brevity. Overall only 46.4% of the scenarios in the parameter space explored led to an acute infection (*Infected - Self Resolving*, *Infected - Untreatable* and *Infected - Treatable*) and were analysed further (table 1), while 53.6% led to a *Failed Infection* and were excluded from further analysis.

**Table 1.** First level classification of simulation scenarios outcomes based on values of key pathogen parameters only.

| simulation scenario<br>outcome | simulation scenarios<br>number | simulation scenarios<br>percentage |
| --- | --- | --- |
| Failed Infection | 44688 | 53.6% |
| Infected - Self Resolving | 32802 | 39.4% |
| Infected - Untreatable | 405 | 0.5% |
| Infected – Treatable | 5454 | 6.5% |

Of the 38661 scenarios leading to acute infections (Table 2), 84% led to a self-resolving infection that did not need any antibiotic course to resolve. 13 − 14% led to an infection which could be successfully treated with a standard fixed duration antibiotic course of 5-10 days, < 1% to a non-treatable infection and 1% to a standard fixed duration course that failed or where the infection relapsed. As detailed above, at the first stage, classifications did not rely on the duration of the fixed antibiotic course, just on the pathogen dynamics parameters. We then applied a next classification stage just to the *Infected-treatable* infections to understand how they were affected by different durations of a fixed antibiotic course (5 vs 7 vs 10 days in Table 2). As explained above, the classification was determined by pathogen abundance at the simulation endpoint. Complete clear-simulation scenario outcome-ance was classified as *Infected - Standard Treatment Succeeds*, while any detectable pathogen was classified as *Infected - Standard Treatment Fails*. The case when an initial decline below the recovery threshold was followed by rebound above the noticeable threshold within 28 days, necessitating further antibiotic treatment was classified as *Infected - Standard Treatment Relapses*. The impact of the different duration of the fixed antibiotic course on this classification was relatively small (Table 2). We therefore chose 7 days as the duration of the standard antibiotic treatment throughout the rest of the simulations.

**Table 2.** Classification of acute infections and number (and percentage) of outcomes for different durations of the standard treatment (5, 7 and 10 days).

| simulation scenario outcome | Fixed 5 days<br>antibiotics | Fixed 7 days<br>antibiotics | Fixed 7 days<br>antibiotics |
| --- | --- | --- | --- |
| Infected - Untreatable | 405 (1%) | 405 (1%) | 405 (1%) |
| Infected - Self Resolving | 32802 (84%) | 32802 (84%) | 32802 (84%) |
| Infected - Standard Treatment Fails | 243 (0.6%) | 162 (0.4%) | 150 (0.4%) |
| Infected - Standard Treatment Relapses | 201 (0.5%) | 144 (0.4%) | 42 (0.1%) |
| Infected - Standard Treatment Succeeds | 5010 (13%) | 5148 (13%) | 5262 (14%) |

### General Model Outcome - parameter sensitivity analysis

Using 7 days as the standard fixed antibiotic treatment duration, we then explored the impact of the different parameters on the infection outcomes (figure 5).

**Figure 5.**
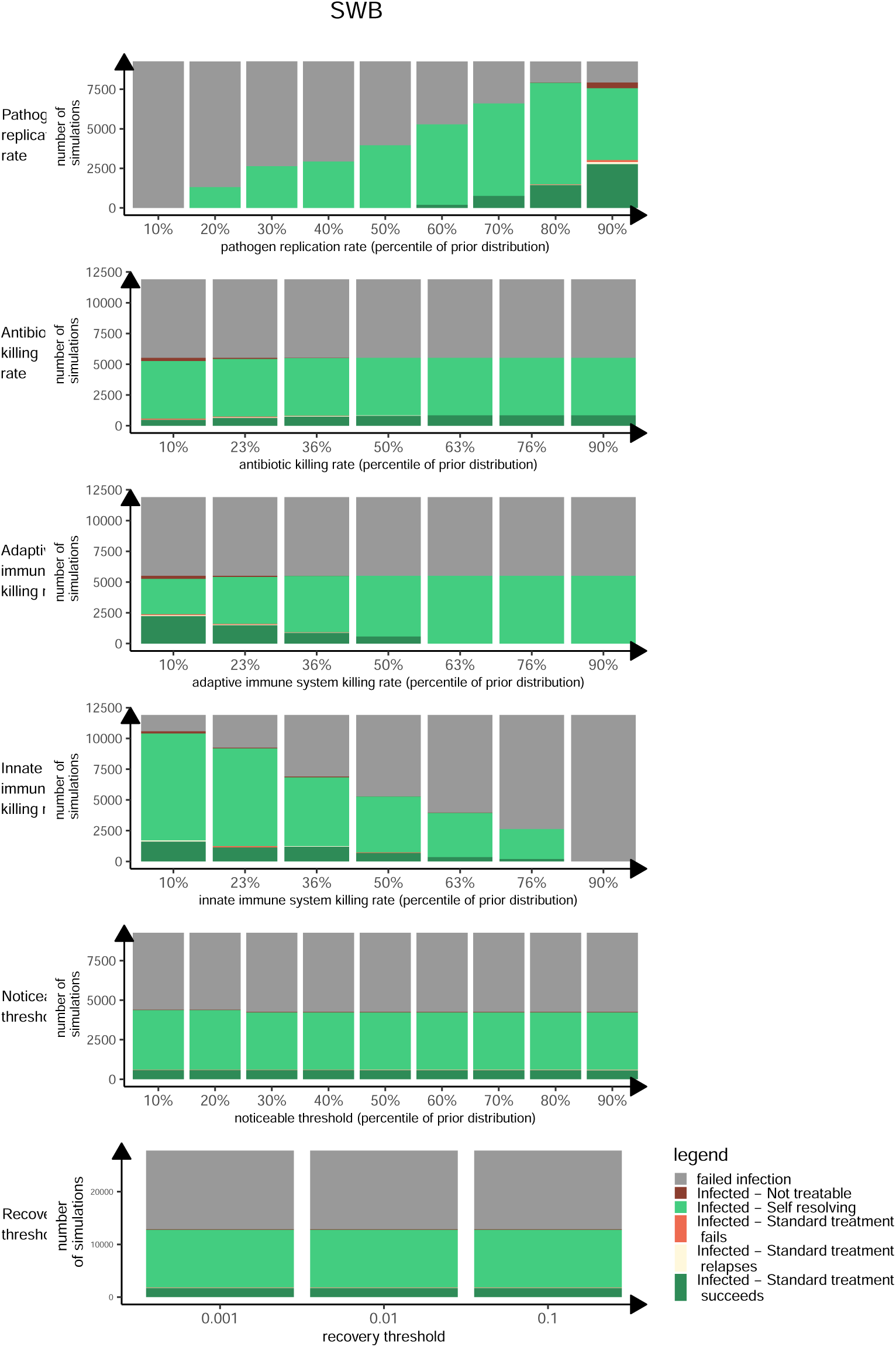
Results of initial simulations to define relevant parameter space for further exploration. Each plot row shows the histogram of outcome for each simulation scenario (detailed on the left) as a function of the value of a different parameter. The parameter values are reported as quantiles of the prior distribution used in the simulations. A detail of the values used can be found in Supplementary. Each simulation is classified as failed infection if an acute infection does not develop. Infected stands for acute infections, that are then classified as untreatable, self resolving, and treatable based on the simulation parameters, and further classified as Standard (7-day) treatment failed, relapsing, or succeeded based on the pathogen abundance at simulation endpoint (28 days).

Panels in figure 5 show the trends in the distribution of the different infection outcomes as a function of the values the parameters (expressed as quantiles in the prior distribution). The pathogen replication rate (figure 5 upper line) is strongly related to infection outcomes, with the number of failed infections decreasing strongly with the pathogen replication rate, as expected, as the higher the quantile in the replication rate, the less likely it is that the pathogen does not reach the noticeable threshold. Higher quantiles in pathogen replication rate were associated with the greatest variability in the acute infection outcomes. The innate immune system killing rate (figure 5 fourth line) was also strongly related to infection outcomes, with the number of failed infections increasing as its value increased. The number of infections successfully treated with a standard 7-day antibiotic course declined with the immune system killing rate, but less sharply than with the increase in the pathogen replication rate. Neither the noticeable threshold nor the recovery threshold (figure 5 fifth and sixth lines) affected any of the infection outcomes, with a fairly constant number of failed infections, and consistent outcomes of acute infections across all parameters. Similarly the antibiotic killing rate (figure 5 second line) did not influence the number of failed infections, but had a small effect on the outcomes of acute infections, with the untreatable infections paradoxically mostly associated with lower rates of antibiotic killing and a trend towards the number of successfully treated infections growing with the antibiotic killing rate. Finally, the adaptive immune system (figure 5 third line) did not affect the number of failed infections, but did affect the outcomes of acute infections with both untreatable infections and infections treatable with standard treatment course declining as the rate increases, that is, as the effective replication rate of the pathogen becomes higher and therefore it becomes less likely for any infection to become acute.

As the innate immune system is always working, the effective pathogen replication rate reflects the difference between the pathogen replication rate and the innate immune system killing rate. This is the only quantity influencing the infection becoming acute, therefore it is not unexpected that these two parameters are the only two showing a strong association with the number of failed infections. The other rates do not affect the number of failed infections, but affect the distribution of the outcomes of acute infections in different ways.

### Commensals

The model block that captures interactions between commensals can be used to explore the development of resistance in a single niche with different values of the niche parameters. Focusing on the scenarios in which the infection is acute and treatable, we modelled the frequency of scenarios ending up in different niche resistance profiles across our parameter space.

The distribution of scenarios among the three niche resistance outcomes (“*baseline resistance*”, “*partially resistant*” and “*fully resistant*”) (Figure 6A, left side) is U-shaped, with around half of the scenarios (63%) leading to a fully resistant niche and around one quarter (22%) to a niche where the resistant population does not exceed the baseline level. The remaining scenarios (15% of the total) led to partially resistant niches.

**Figure 6.**
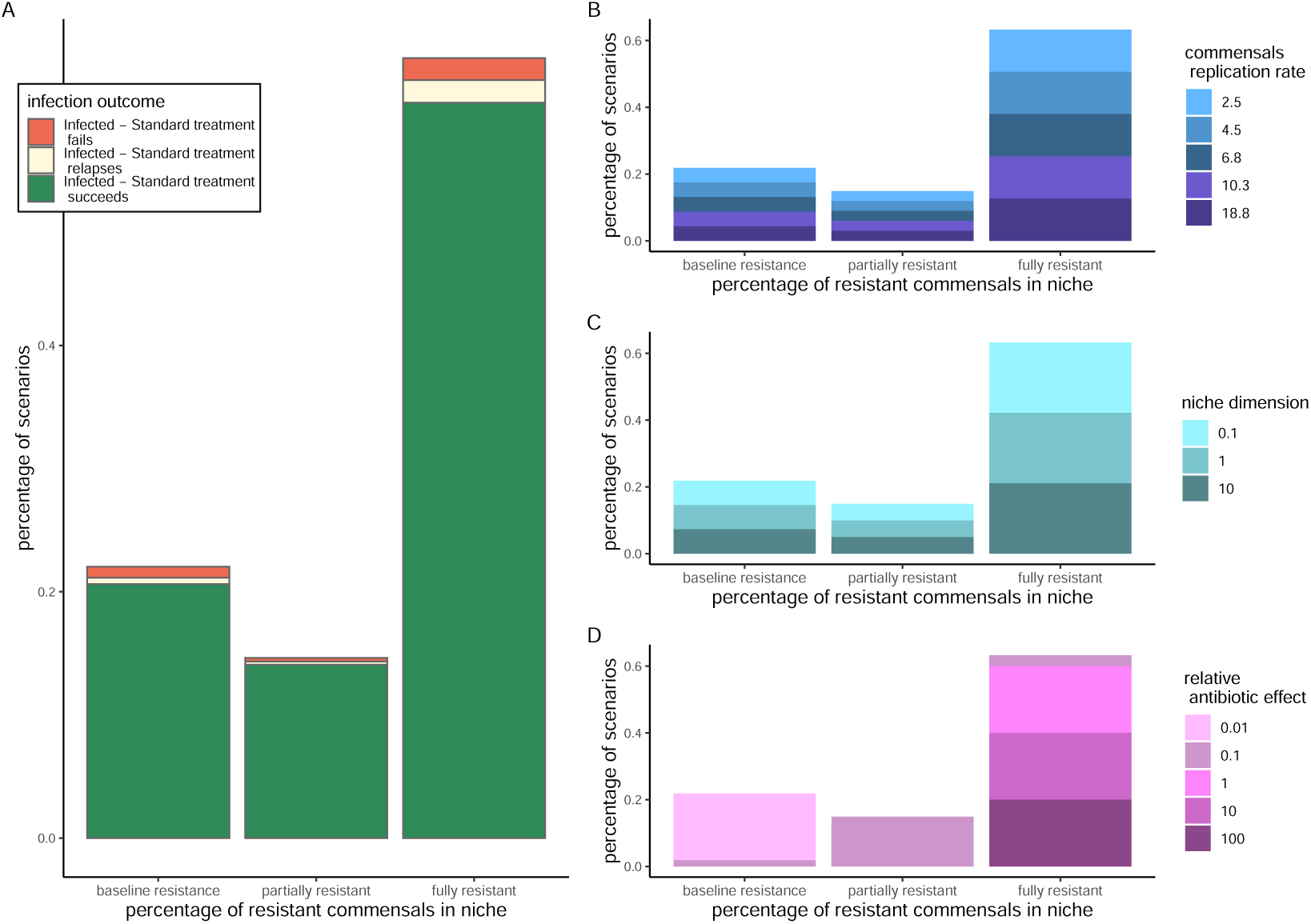
Characterization of determinants of resistance prevalence in niches. **A:** Distribution of percentage of resistant commensals in the niche as a function of treatment outcomes in the standard treatment scenarios. **B-C-D:** Distribution of percentage of resistant commensals in the niche as a function of the different parameters affecting commensals dynamics: commensals replication rates (B), commensals relative niche dimension (C), relative effect of the antibiotic killing rate in the niche (D).

Figure 6 B-C-D (right side) shows the percentage of commensals in the niche as a function of the values of the parameters determining the commensals dynamics: commensals replication rate, relative niche dimension and relative antibiotic effect. The distribution of scenarios in each outcome is evenly split across parameters in the commensals replication rate and niche size, while for relative antibiotic effect the stronger the effect of the antibiotic, the more likely it is that the niche becomes fully resistant.

### Application of the model to SWB

We applied this model to study the potential impact of a Stop When Better (SWB) approach to prescribing. We explored how much shorter the treatment would be in the case where, instead of a fixed duration, an antibiotic treatment was stopped when the patient was “*feeling better*”, and what would happen where “*feeling better*” did not correspond to sufficient recovery from the infection, hence triggering a relapse of the infection and another subsequent antibiotic course. In our simulations, the patient “*feeling better*” is taken to be the point at which the pathogen abundance falls below a predefined “*recovery threshold*”. The recovery threshold is defined as a multiplicative constant strictly smaller than 1 applied to the noticeable threshold, assuming that the recovery threshold is below the noticeable threshold.

We simulated the outcome of the infection in the SWB approach and compared the length of antibiotic exposure using the standard course and the “stopping when better” approach. We limited our exploration to acute infections that can be treated with a standard treatment as these are the ones with more potential for implementing a stop when better regimen: we know already that the standard treatment works, and ask the question whether a shorter treatment could adequately treat the infection while at the same time ensuring shorter exposition in the commensals? We therefore excluded from further analysis all the simulations that in the standard treatment regimen had been classified as “*failed infection*” (44688 scenarios), “*infected - untreatable*” (405 scenarios), “*infected – self-resolving*” (32802 scenarios). We analysed the remaining 5454 scenarios, originally classified as “*infected - treatable*”. These scenarios account for 6.5% of the initial parameter combinations (table 1).

### Alternative implementations of SWB

We considered two different implementations of SWB. In the simplest one, the antibiotic is stopped as soon as the pathogen abundance hits the “*recovery threshold*”. This implementation, called **SWB** for brevity, may be too simplistic as it assumes that antibiotic exposure occurs at a constant rate over time, when in reality antibiotics are administered periodically at discrete time intervals (either one or multiple doses spread evenly throughout an individual’s day), and that a patient is immediately aware of abundance reaching an (arbitrary) threshold, rather than symptoms improving. Therefore, to make the model more realistic, we considered a second implementation, where the antibiotic is stopped, and its effect ceases, a fixed amount of time (set to 1 day) after the pathogen abundance hits the recovery threshold. This time accounts for the patient realising they feel better, stopping taking the prescribed antibiotic and the residual pharmacokinetic impact of the last pill taken. We call this alternative implementation **SWB+1**. An example of the pathogen dynamics under the SWB+1 scenario is shown in figure 2 panel B. An example of how it affects the different simulation outcomes classification is shown in the right column in figure 4 (which uses the same parameters as in the left column). Particularly striking is the effect on the “Infected-treatable” (figure 4 bottom row) scenario, where relapses in the SWB scenario get fully treated in the SWB+1 scenario.

### Comparison of Standard Treatment and SWB

We started by comparing the standard treatment duration and SWB in all the “*infected - treatable*” scenarios, including scenarios where the standard treatment failed. The main focus of the comparison was the length of the primary antibiotic course, but we also considered whether or not there was a relapse in infection, as a relapse triggers an entirely new subsequent standard antibiotic course, potentially making the SWB approach longer than the standard treatment for an individual and even on average at the population level.

Each SWB scenario was classified in terms of the comparison between the primary cycle length as “*shorter*”, “*equivalent*” (if the difference in course length was within a day), “*longer*” or “*failure to treat*” as compared to the standard 7-day course with the same parameters (see figure 4 bottom line left panel dotted line for an example of relapsing infection in the SWB scenario). To all these classifications, the suffix “*-relapse*” was added if the infection relapsed, triggering a subsequent antibiotic course. In table 3 we summarise the outcomes of the simulations in the SWB scenario according to the outcome from the standard course.

**Table 3.**
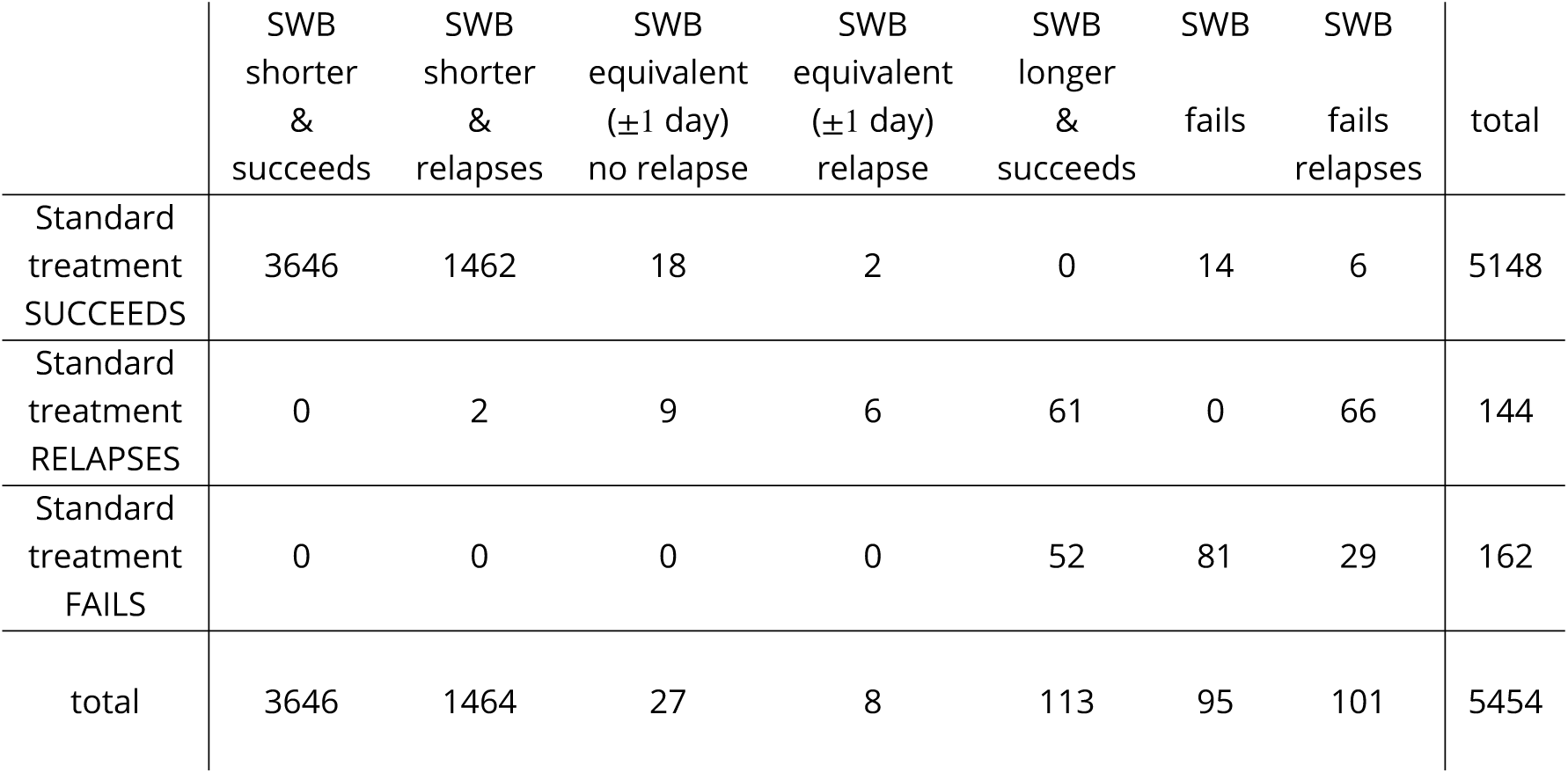
SWB outcomes versus the standard course.

Two thirds of all the scenarios led to a shorter successful treatment in the SWB approach (3646∕5454; 71% of all scenarios where standard treatment succeeded) but a further 27% of all scenarios led to a shorter primary treatment with SWB but with the infection subsequently relapsing, leading to longer antibiotic exposure overall (1464∕5454). This happened across all ranges of the shorter SWB duration (Figure 7 top row). The remaining 6% of all scenarios led to a range of outcomes detailed in table 3, including successful longer primary SWB treatment where standard treatment relapsed or failed (113∕5454, 3%; 37% of scenarios where the standard treatment failed/relapsed) and failure of both approaches.

**Figure 7.**
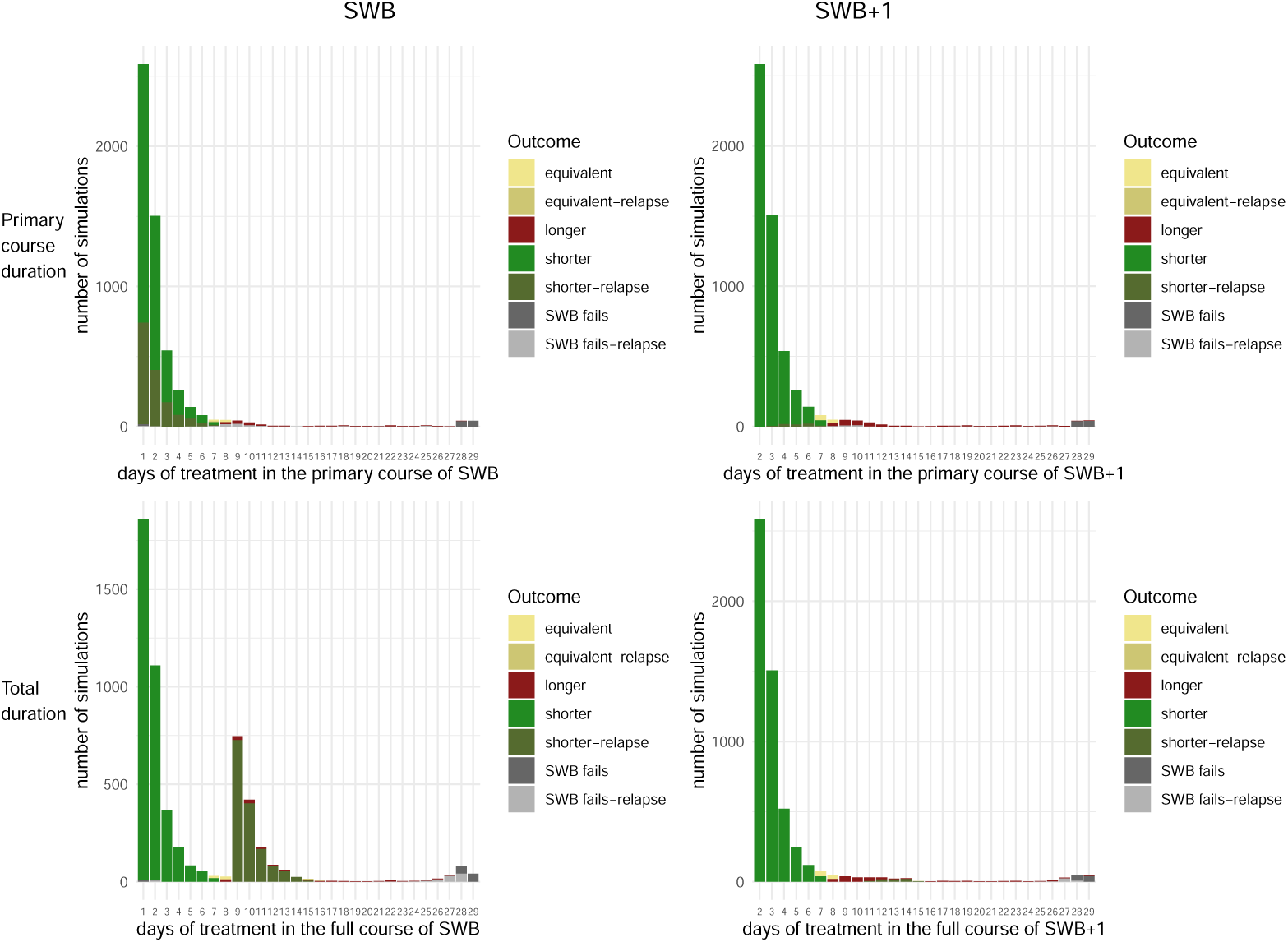
Duration of the antibiotic course. **Top-Left:** Duration of the primary course in the SWB scenario. **Top-Right:** Duration of the primary course in the SWB+1 scenario. **Bottom-Left:** Duration of the total antibiotic course (primary+relapse) in the SWB scenario. **Bottom-Right:** Duration of the total antibiotic course (primary+relapse) in the SWB+1 scenario. Colors refer to the comparison with the standard treatment scenario (7 days).

In figure 7 top Left panel we show the duration of the primary SWB/SWB+1 course in days (to the nearest day). The number of infections decayed exponentially as the primary course duration increased.

The impact of the different simulation parameters on determining whether the SWB approach was overall shorter than the standard antibiotic course is detailed in the left column in figure 8. Both the recovery threshold and the noticeable thresholds show clear trends in terms of higher values being associated with shorter SWB primary courses that relapse. The closer the recovery threshold is to the noticeable threshold, the more likely is relapse. The farther the recovery threshold is from the noticeable threshold the more likely it is that the SWB primary course is longer than the fixed 7 day duration. In absolute values, the lower the noticeable threshold, the less likely a relapse.

**Figure 8.**
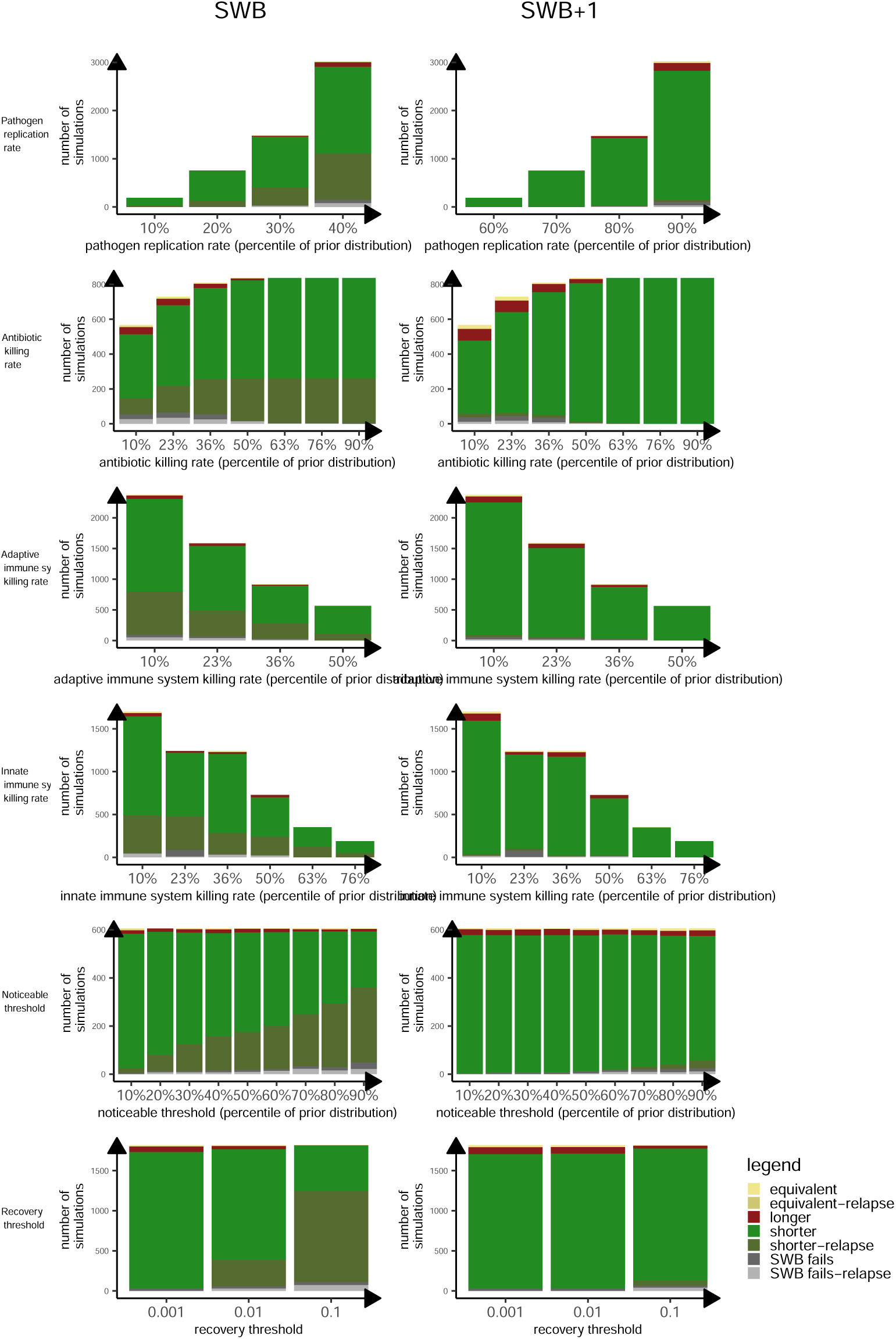
Distributions of treatment outcomes for different values of the parameters in treatable infections managed under SWB or SWB+1 approaches. From top: pathogen replication rate, antibiotic killing rate, adaptive immune system killing rate, innate immune system killing rate, noticeable threshold and recovery threshold. Left column: SWB. Right column: SWB+1. Color code same as figure 7.

### Comparison of Standard course and SWB+1

The SWB+1 regimen was more successful in delivering effective treatment with no relapse compared to both standard course durations and SWB (see table 4). In 92% of the scenarios the infection was treated using SWB+1 with a shorter primary course and no relapse (5010∕5454; 97% of scenarios where standard treatment succeeded), with only 1% of scenarios with a shorter SWB+1 course relapsing (60∕5454). SWB+1 also increased the number of successful longer primary SWB treatments where standard treatment relapsed or failed (183∕5454, 3%; 60% of scenarios where the standard treatment failed/relapsed vs 37% with SWB). Interestingly, the SWB+1 regimen also reduced the number of treatments that failed: 81 vs 95 with SWB.

**Table 4.** SWB+1 outcomes versus the standard course.

|  | SWB+1<br>shorter | SWB+1<br>shorter<br>relapse | SWB+1<br>equivalent | SWB+1<br>equivalent<br>relapse | SWB+1<br>longer<br>succeeds | SWB+1<br>fails | SWB+1<br>fails<br>relapses | total |
| --- | --- | --- | --- | --- | --- | --- | --- | --- |
| Standard<br>treatment<br>SUCCEEDS | 5010 | 60 | 58 | 0 | 20 | 0 | 0 | 5148 |
| Standard<br>treatment<br>RELAPSES | 0 | 0 | 1 | 1 | 115 | 0 | 13 | 144 |
| Standard<br>treatment<br>FAILS | 0 | 0 | 0 | 0 | 68 | 81 | 27 | 162 |
| total | 5010 | 60 | 59 | 1 | 203 | 81 | 40 | 5454 |

The length of the primary course was, as expected, approximately one day longer with SWB+1 (figure 7 upper right panel), but with clear reduction in relapsing infections considering total antibiotic exposure (figure 7 lower right panel).

When examining the role of the different simulation parameters in the treatment outcome (fig 8 right column) the impact of noticeable and recovery rate was similar to SWB but substantially attenuated.

### Resistance development in the niche

The length of the exposure to antibiotics plays a strong role in resistance development in commensals, but other niche-specific parameters play a role too, in particular the relative antibiotic killing rate and the commensals replication rate. We therefore simulated the commensal niches, restricting for simplicity to scenarios that led to a shorter primary course with no relapse using SWB+1 compared with the fixed 7 day standard course. As above, we classified the abundance of resistance in each niche according three possible outcomes: “*baseline resistance*”, “*fully resistant*” and “*partially resistant*”.

From the perspective of generating resistance in the commensal niche, the preferred prescription approach would be the one that maximised the number of niches classified as baseline resistance, as these niches do not significantly develop resistance, and minimised the number of niches that become fully resistant.

Figure 9 shows the proportion of scenarios leading to the different resistance outcomes as a function of the prescription approach (x axis) and explicitly separating the cases where SWB relapses from the cases where SWB does not relapse. Note that we choose the scenarios in these simulations as those in which SWB+1 does succeed and does not relapse. So we are comparing a successful SWB+1 scenario to a SWB that does not relapse (figure 9 bottom line) or a SWB scenario that does relapse (figure 9 top line).

**Figure 9.**
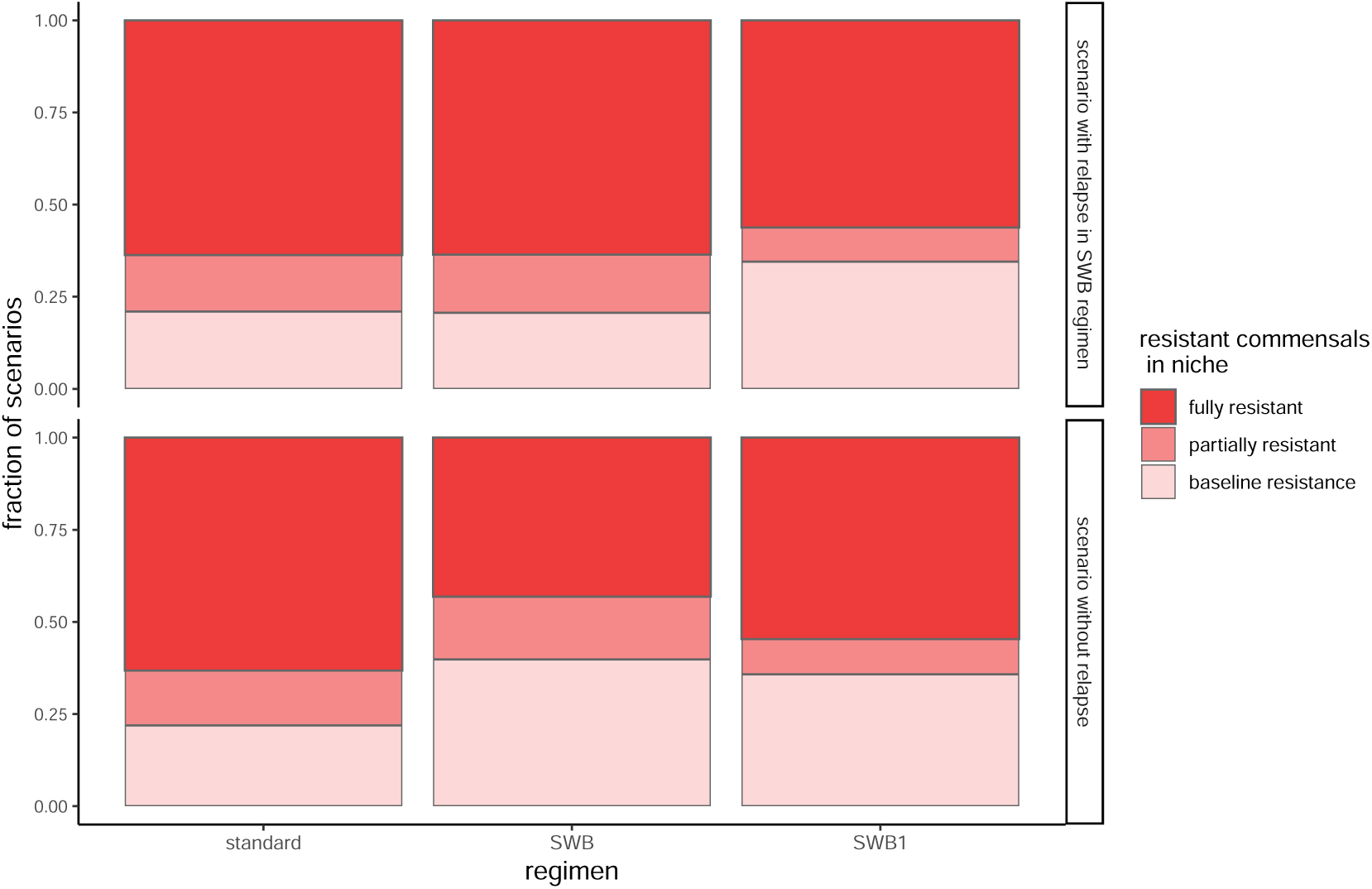
Proportion of niches with different resistance outcomes. Resistacne outcomes in niches are stratified by prescribing approach and whether the primary course leads to relapse in the SWB scenario. In dark red we show the abundance of fully resistant niches, in pink the abundance of partially resistant niches, in light pink the abundance of baseline resistant scenarios. The columns are normalised to the total number of simulations which relapse (**upper panel**) or do not relapse (**lower panel**) in the SWB approach.

In case of SWB with relapse, the SWB+1 regimen generates less resistance overall, i.e. more “*baseline resistant*” scenarios and less “*fully resistant*” scenarios. In fact, in the case of SWB relapse, the SWB+1 regimen (which lasts one day longer than the SWB approach but does not relapse) generates 35% of scenarios with “*baseline resistance*” (35985∕104325) versus 20% in the SWB approach (21540∕104325). It also generates less “*fully resistant*” scenarios: 56% in the SWB+1 vs 63% in SWB. So overall the SWB+1 scenario generates fewer resistant niches.

Figure 9 shows clearly that the standard treatment regimen generates more resistance compared to the other prescribing regimes: fewer “*baseline resistance*” niches both when the infection relapses (21% of the scenarios; 21900∕104325 SWB relapsing scenarios) and does not relapse in the SWB scenarios (22%; 59550∕271425 SWB non-relapsing scenarios). In both the relapsing and nonrelapsing scenarios the standard 7-day course leads to a similar proportion of fully resistant niches: 63% (171540∕271425) SWB non-relapsing scenarios and 64% (66465∕104325) SWB relapsing scenarios.

## Discussion

We have presented a model describing the within patient dynamics pathogen during an infection and its subsequent antibiotic treatment. The model describes the dynamics at the level of the interactions between the pathogen, the immune system and the antibiotic treatment, leading to the infection becoming acute, self-resolving without needing antibiotics, needing antibiotics for treatment, with a primary course either being successful or failing initially or completely. The model also describes how the antibiotic treatment acts on the commensal niches, helping us quantify how much resistance can be generated from an antibiotic course. We applied the model to study the possible outcomes of implementing a “*stop when better*” approach to prescribing, which would instruct the patients to stop the antibiotic treatment when they “*feel better*”. We studied two different implementations: one in which the antibiotic was stopped immediately when the pathogen abundance reached the “*recovery threshold*” (called SWB), which was our proxy for symptoms improving, and another when the antibiotic was stopped one day after the pathogen abundance reached the “*recovery threshold*” (called SWB+1). Our simulations showed that the SWB+1 approach led to shorter overall antibiotic exposure and less relapse compared to SWB and that both led to more shorter successful treatments compared to the standard course. This difference in treatment duration was reflected in lower resistance development in commensal niches.

This model is a mathematical description of the processes happening within the patient during an infection and its subsequent treatment. There are several gaps that prevent the direct mapping of this model and its results into clinical practice. One is the parametrization of the model: many of the parameters, including the pathogen replication rate did not have biological measurements, so we had to find a way to parametrise them. Many studies provide a reliable measure for the pathogen replication rate in vitro, but there is no direct measure in vivo, except maybe in very specific and unrealistic conditions (e.g. time it takes for an uncolonised mouse to become colonised). We assumed in our simulations a pathogen replication rate normally distributed between one hour and one day, which seems realistic for the community-acquired infections we are modeling. Still, the pathogen replication rate is a well defined and well studied quantity as compared to the killing rate of the adaptive and innate immune systems or the antibiotic killing rate in different niches. The “*noticeable threshold*” and “*recovery threshold*” are debatable both from the modeling and measurement perspectives. While the idea of the pathogen abundance being relevant in the course of the infection can be realistic, having such an immediate effect of stopping antibiotics is plausibly just a modeling construct; hence our introduction of the SWB+1 approach. Despite these limitations, this model was created as a proof of concept for the entire process of pathogen infection and treatment. As such none of its parameters should be taken literally, but rather results used to identify the types of pathogens where a SWB+1 approach might have benefits. In particular our analysis points as the best case scenario to implement a “*stop when better*” approach to pathogens with a “*fast*” replication rate and antibiotics with a “*high*” killing rate in patients that complain early (low noticeable threshold) and do not rush in feeling better (low recovery threshold).

There are several key evidence gaps around applying a “*stop when better*” approach in clinical practice that need to be investigated. While the meaning of “*feeling better*” seems intuitive, giving an operational definition that can be used by both patients and prescribers is an obstacle to implementing such an approach to prescribing ***Borek et al. (2024)***. Also both patients and prescribers have concerns around its safety and clinical trials are needed to determine the impact of such an approach for both clearing infections and reducing the risk of resistance in the microbiome. Further, it has been demonstrated that patients may personally decide to stop taking antibiotics despite guidance to the contrary, i.e. are already following a SWB(+1) approach and potentially gaining its benefits. We have not considered this in our model.

Ideal treatment length is likely impacted by a number of individual-level factors such as age, gender, comorbidities and others, therefore not every patient may be suitable for a “*stop when better*” approach. Nonetheless the duration of prescribing is significantly associated with the risk of resistance and strategies to reduce antibiotic use should be further studied. Another approach to reducing prescribing might be research into diagnostic tests that could determine the most appropriate treatment length for an infection. A model such as this one is a useful starting point for demonstrating the need for these studies. Finally, we have not considered the impact of inappropriately prescribing antibiotics in this case but there is a high potential for impact in this area.

While this model is far from providing any insight directly applicable to clinical practice, it is a good proof of concept tool to guide the discussion on changing our approach to antibiotic prescribing. Whether we need to develop prescription duration tailored on the patient or on the pathogen causing the infection, there is still a lot that we can do to optimise our prescribing strategies in order to maximise our treating power while minimising antibiotic resistance developments in commensals (and hence future comorbidities).

## Data Availability

All data used in the manuscript comes from simulations. The code for those simulations is available on github.

## Acknowledgments

This study was funded by the National Institute for Health and Care Research (NIHR) Health Protection Research Unit in Healthcare Associated Infections and Antimicrobial Resistance at the University of Oxford in partnership with the UK Health Security Agency (UKHSA) [NIHR200915]. ASW. is an NIHR Senior Investigator and is also supported by the NIHR Oxford Biomedical Research Centre. KBP. is also supported by the Medical Research Foundation (MRF-160-0017-ELP-POUW-C0909). The views expressed in this publication are those of the authors and not necessarily those of the NHS, the NIHR, the Department of Health or the UKHSA.

## Appendix 1

### Analytical description of the model

The model is described by a set of ordinary differential equations describing the pathogen and the commensals dynamics. The equations are mostly deterministic with stochasticity only affecting the probability of getting a resistant colony from the environment.

### Equations

The equations for the abundance of pathogen and for the abundance of susceptibles and commensals in each niche are as follows:

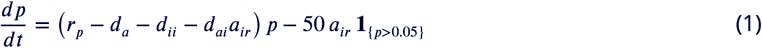

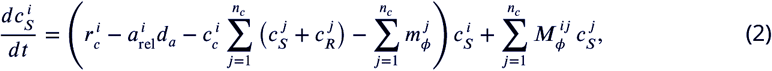

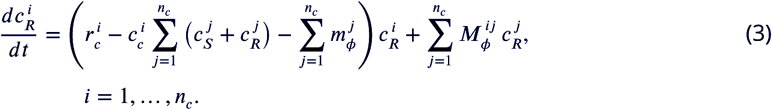

The meaning of the variables and parameters is detailed in the next two paragraphs and tables.

### Variables & Parameters

The model is written as a set of ordinary differential equations that describe the evolution of three main variables: the pathogen abundance, the the abundance of susceptible and commensal bacteria in each niche. So the total number of variables is (2 x i) + 1 where i is the number of niches that are simulated (see table 4). The parameters are detailed in table M2.

**Table 1.** The variables in the equations, their description and their initial value.

| Variable | Description | initial value |
| --- | --- | --- |
| $p$ | pathogen abundance | $p_0$ |
| $c_S^i$ | abundance of susceptible commensals in niche $i$ | $c_{S0}$ |
| $c_R^i$ | abundance of resistant commensals in niche $i$ | $0.01c_{S0}$ |

**Table 2.** All the parameters used in the equations and their description. The priors and simulated values of the parameters are given in table M5 and figure MF1.

| Parameter | Description |
| --- | --- |
| $r_p$ | pathogen replication rate |
| $d_a$ | antibiotic killing rate |
| $d_{ai}$ | adaptive immune system killing rate |
| $d_{ii}$ | innate immune system killing rate |
| $p_n$ | noticeable threshold (pathogen abundance) |
| $p_r$ | recovery threshold (percentage of pathogen abundance) |
| $r_c^i$ | commensals replication rate (in each niche) |
| $c_c^i$ | competition rate in the niche ($c_c = r_c k$) |
| $a_{rel}^i$ | relative killing rate of the antibiotic in the niche (pure number) |
| $m_\phi^i$ | rate of importation of new bacteria in niche i |
| $M_\phi^{ij}$ | rate of bacterial exchange between niches i and j |

#### Classification of the simulations

First stage classification is based only on the values of the simulation parameters.

**Table 3.** First level classification based only on the values of the parameters.

| Classification | Condition |
| --- | --- |
| **Acute infection** | $p > p_n$<br>*within 14 days* |
| Infected<br>**Self-Resolving** | $p > p_n$ &<br>$r_p - d_{ii} - d_{ai} < 0$ |
| Infected<br>**Untreatable** | $p > p_n$ &<br>$r_p - d_{ii} - d_{ai} - d_{au} > 0$ |
| Infected<br>**Treatable** | $p > p_n$ &<br>$0 < r_p - d_{ii} - d_{ai} < d_{au}$ |

### Implementation of the model

The model was implemented in R using the library *deSolve* (cite)

### Details of the programs

The core of the SWB simulation is implemented in the file funcs-ode-v8.0Final.R, which simulates both the standard and the SWB scenario. The SWB+1 scenario is implemented in the file funcs-ode-v8+2Final.R together with the standard scenario. In both files there is a function called simul that takes as an input a named vector with the values of all the parameters and produces a matrix with the status of the variables and the parameters at each time point. The simul function itself is developed in three main steps. As the pathogen is expected to be arriving in an otherwise healthy person, the first step of the simulations is finding the equilibrium state in the commensal niches, given the possibility that the niches exchange bacteria and the fact that each niche has its own size and replication rate. This step is not relevant when we are dealing with a single niche as the equilibrium state of the single niche is determined by the parameters defining the niche’s dynamic. The second step is determining the initial abundance of resistant bacteria in each niche given the (Stochastic) probability of importing resistant bacteria from the environment. Finally, the third part of the simulation is the one simulating the pathogen dynamics alongside the niches dynamics.

### Switches

The model equations are made of distinct terms that are conditionally activated or deactivated based on threshold-driven discrete variables. For example, the term accounting for the first antibiotic treatment is proportional to *d_a_* ⋅ *au*, with *d_a_* being the antibiotic killing rate. *au* is a binary variable set to 0 at the beginning of the simulations, making the term nil. When the pathogen abundance reaches *p_n_* the parameter *au* is switched on (*au* = 1), making the term contribute to the pathogen dynamics. In the SWB simulations *au* is switched off (*au* = 0) upon the pathogen abundance reaching the recovery threshold (*p* = *p_n_*). In the SWB+1 scenario, reaching the recovery threshold triggers a timer that stops the antibiotic in one day (*au* = 0).

**Table 4.** Switch parameters used in the equations, their description and their value at the beginning of the simulations.

| Variable | Description | initial value |
| --- | --- | --- |
| *my – treatment* | switch between the standard and the SWB (SWB+1) prescription regimen | 0/1 |
| *timer* | track the duration of the antibiotic treatment | –1 |
| *air – timer* | switch on the adaptive immune system | –1 |
| *au* | binary variable to track the first antibiotic treatment | 0 |
| *au2* | binary variable to track the second antibiotic treatment (relapse) | 0 |
| *air* | adaptive immune system on | 0 |
| *lag* | lag to adaptive immune system to work | 0.1 days |
| *stop – timer* | timer to track the one day lag in stopping the antibiotic treatment in the SWB+1 scenario | –1 |

**Table 5.**
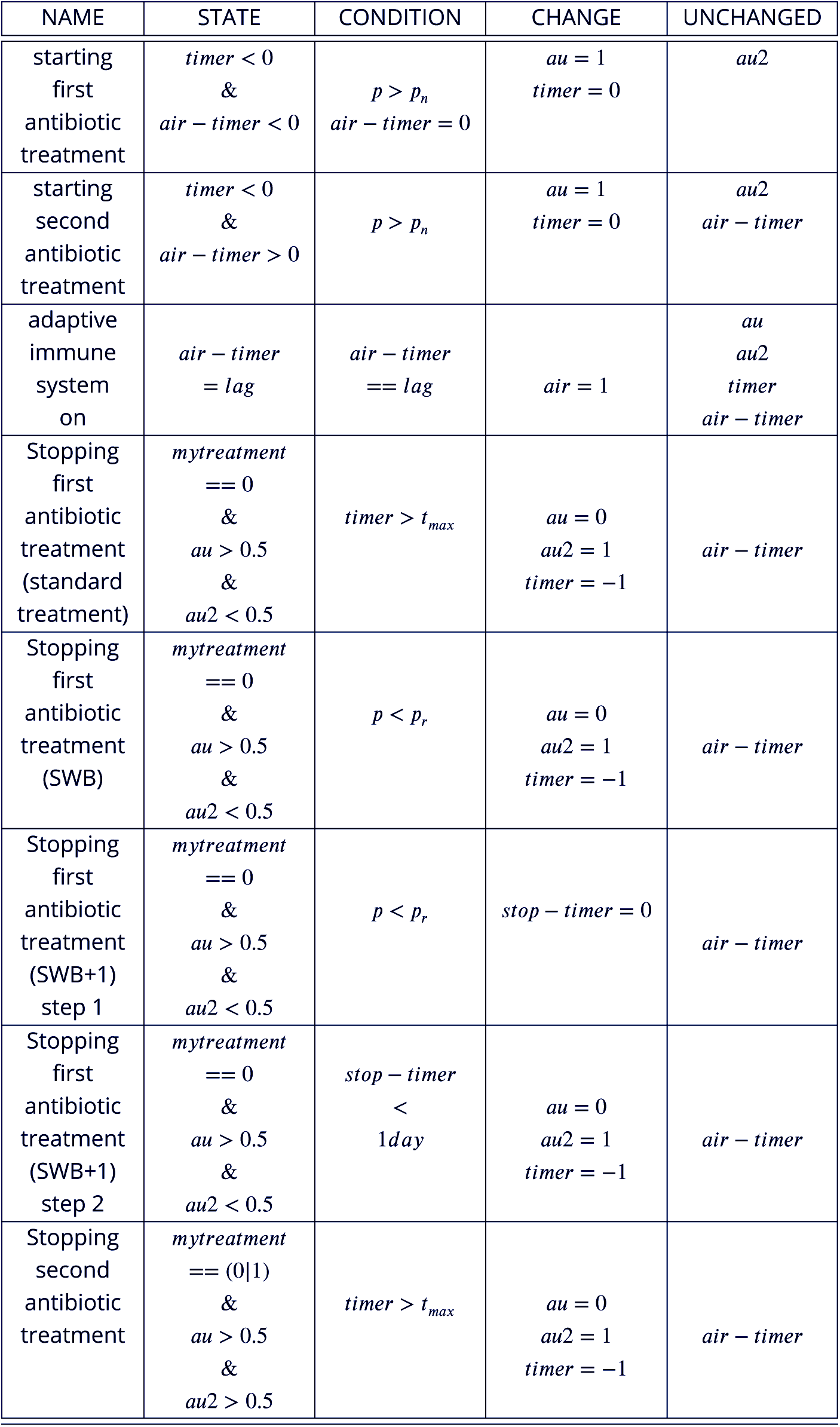
Switch values used in the equations to account for the different simulated situations. For each switch we provide a brief description of its function, the state of the variables needed for the condition to be checked, the condition that triggers the switch, the changes in the variables that the switch triggers and the variables that are not affected by the switch.

### Choice of the parameters for the simulations

The simulations were realised on a latin hypercube. Since there is no direct data to inform the model, we had to determine sensible priors for the parameter

**Figure 1.**
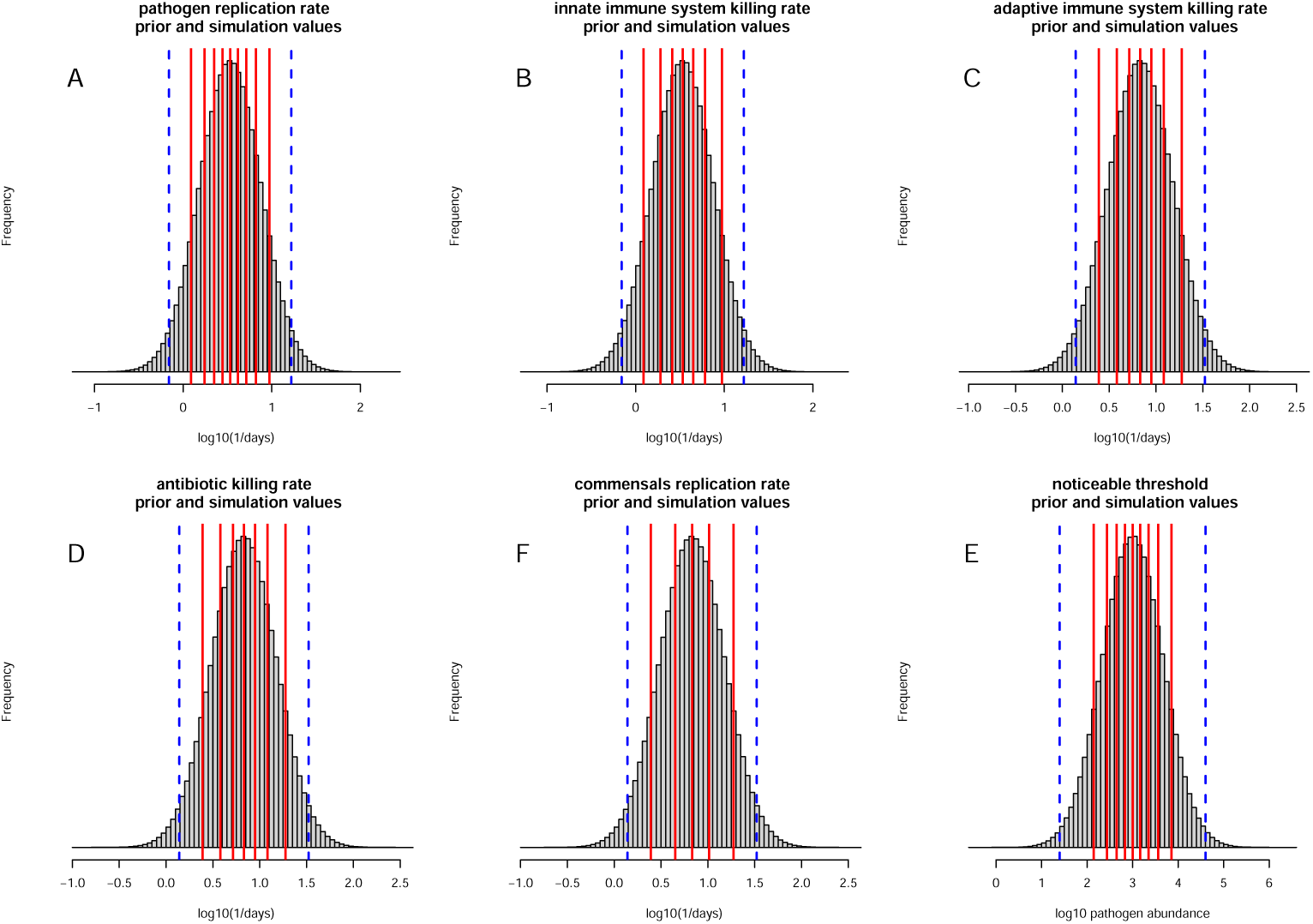
Priors for the simulation parameters. Priors for all the rates (pathogen replication rate, innate immune system killing rate, adaptive immune system killing rate, antibiotic killing rate, commensals replication rate) and noticeable threshold. The all are a normal distribution where the dashed blue lines denote the 99% interval. The red lines denote the quantiles of the distribution used in the simulations. Their values are given in table. **A. pathogen replication rate:** duplication rate between an hour and a day; **B. innate immune system replication rate:** rate between an hour and a day. **C. adaptive immune system killing rate:** rate between half an hour and half a day; **D. antibiotic killing rate:** rate between half an hour and half a day; **E. commensals replication rate:** duplication rate between half an hour and half a day; **F noticeable threshold:** prior for the noticeable threshold value, expressed as *log*_10_ of pathogen abundance. It is a normal distribution centered on 1000 pathogens and with 99% interval between 25 and 40.000.

All the rates were extracted from a normal distribution with 99% interval between a replication rate of half an hour and a replication rate of half a day (figure 1 panels A, B, C). This seems reasonable considering the kind of infections that we want to study. The prior distribution for the noticeable threshold pn is a normal distribution centered on a colony of 1000 individuals and with the 99% interval ranging from 25 individuals to 40000 individuals (figure 1 panel D). The prior for the recovery threshold pr (which is a multiplicative factor to the noticeable threshold) is a flat prior between 0.001 and 0.1 and the values for the simulation values have been chosen to be equispaced on logarithmic scale. The prior for the niche carrying capacity is flat between 0.1 and 10 and the prior for the relative strength of the antibiotic killing rate on commensals niches is flat between 0.01 and 100.

The values of the parameters are detailed in table 9 Value of the parameters and in figure 10 where the red lines in the prior distribution denote the values used in the simulations. The latin hypercube simulating the pathogen scenarios is made by 9 ⋅ 7 ⋅ 7 ⋅ 7 ⋅ 9 ⋅ 3 = 83349 combinations of the parameters.

The parameters values used to create the latin hypercube to simulate the commensals dynamics is given in the script generateLHPathogen. . . R. The additional parameters from the commensals parameters are in generateLHCommensals. . . R. The values of the parameters are detailed in table M5 Value of the parameters and in figure 10 where the red lines in the prior distribution denote the values used in the simulations.

**Table 6.** values of the parameters used in the simulations. Where the value was chosen as a quantile of the prior distribution, the quantile value is given in parenthesis.

| Parameter | Values (quantiles) | total |
| --- | --- | --- |
| $r_p$ | 1.23 (10%), 1.74 (20%), 2.23 (30%), 2.77 (40%), 3.39 (50%),<br>4.15 (60%), 5.15 (70%), 5.15 (70%), 6.23 (80%), 9.40 (90%) | 9 |
| $d_a$ | 2.45 (10%), 3.81 (23%), 5.18 (36%), 6.79 (50%),<br>8.91 (63%), 12.11 (76%), 18.80 (90%) | 7 |
| $d_{ai}$ | 2.45 (10%), 3.81 (23%), 5.18 (36%), 6.79 (50%),<br>8.91 (63%), 12.11 (76%), 18.80 (90%) | 7 |
| $d_{ii}$ | 1.23 (10%), 1.90 (23%), 2.59 (36%), 3.39 (50%),<br>4.45 (63%), 6.05 (76%), 9.40 (90%) | 7 |
| $p_n$ | 139 (10%), 275 (20%), 447 (30%), 677 (40%), 999 (50%),<br>1475 (60%), 2237 (70%), 3636 (80%), 7142 (90%) | 9 |
| $p_r$ | 0.001, 0.01, 0.1 (· pathogen abundance) | 3 |
| $r_c^i$ | 2.45 (10%), 4.47 (30%), 6.79 (50%), 10.30 (70%), 18.80 (90%) | 5 |
| $k$ | 0.1, 1, 10 | 3 |
| $a_{rel}^i$ | 0.01, 0.1, 1, 10, 100 | 5 |

### Pathogen simulations

The first set of simulations was realised on a latin hypercube covering 9 values each for the pathogen replication rate rp and the noticeable threshold pn, 7 values each for the antibiotic killing rate da and the innate and adaptive immune system (dii and dai), and 3 values for the multiplicative recovery threshold pr. All these values produce 83349 combinations of parameters which were simulated for 3 different durations of primary antibiotic cycle. The code doing the simulation and a preliminary analysis of the simulation (to have a storable outcome) is *newpar*_7_.*R*. This script takes as input a vector with all the values of the parameters, generates the simulation and then processes the output matrix to store the relevant information in one single line.

The output of this script is stored in the file *log*2*_P_ at*ℎ_7_.*txt*.

This file is then processed using the script *New_F_ ig*1_7_*days*.*R* to extract the information and the plots described in Results. **Cite the script multiplot** For the SWB+1 scenario the simulation script is *newpar*_7_ +1.*R* that produces the output *log*2*_P_ at*ℎ_7_ +2.*txt* that is then elaborated by the script *New_F_ ig*1_7_*days*.*R*.

The scripts for generating the plots describing the dynamics for each set of parameters from the output matrix of the simul function are in functions in the script niceplot.R

### Commensals simulations

The parameters describing the commensals dynamics are the commensals replication rate rc, the niche carrying capacity k and the relative strength of the antibiotic in the niche. The prior and simulation values for these parameters is described above.

We used 5 values for the commensals replication rate, 3 values for the niche carrying capacity and 5 values for the relative strength of the antibiotic, which lead to 75 commensals scenarios. All the 75 commensals scenarios were simulated for each of the 5010 pathogen scenarios that led to a shorter treatment (compared to the standard one) in the SWB+1 approach. Overall we simulated the dynamics in 375750 commensals scenarios (files *commensalsLH / commensals_k_.csv*). The script to generate the scenarios is *generateLHCommensals1Niche.R* We simulated the commensals dynamics in the three prescription regimens: the standard regimen, the SWB approach (*newpar*_7_0_*C*_*ommensals_k_.R*) and the SWB+1 regimen (*newpar*_7_ + 1_*C*_*ommensals_k_.R*)

